# Efficacy of moist heat decontamination against various pathogens for the reuse of N95 respirators in the COVID-19 emergency

**DOI:** 10.1101/2020.05.13.20100651

**Authors:** Ebru Oral, Keith K Wannomae, Dmitry Gil, Rachel Connolly, Joseph A Gardecki, Hui Min Leung, Orhun K Muratoglu, Amy Tsurumi, Laurence Rahme, Tareq Jaber, Cassidy Collins, Julian Gjore, Amanda Budzilowicz

## Abstract

Decontamination of N95 respirators has become critical to alleviate PPE shortages for healthcare workers in the current COVID-19 emergency. The factors that are considered for the effective reuse of these masks are the fit, filter efficiency and decontamination/disinfection level both for SARS-CoV2, which is the causative virus for COVID-19, and for other organisms of concern in the hospital environment such as Staphylococcus aureus or Clostridium difficile.

The efficacy of inactivation or eradication against various pathogens should be evaluated thoroughly to understand the level of afforded disinfection. Methods commonly used in the sterilization of medical devices such as ionizing radiation, vaporized hydrogen peroxide, and ethylene oxide can provide a high level of disinfection, defined as a 6 log_10_ reduction, against bacterial spores, considered the most resistant microorganisms. CDC guidance on the decontamination and reuse of N95s also includes the use of moist heat (60°C, 80% relative humidity, 15-30 min) as a possible recommendation based on literature showing preservation of fit efficiency and inactivation of H1N1 on spiked masks.

Here, we explored the efficacy of using moist heat under these conditions as a decontamination method for an N95 respirator (3M 1860S, St. Paul, MN) against various pathogens with different resistance; enveloped RNA viruses, Gram (+/-) bacteria, and non-enveloped viruses.

## Background

The efficacy of inactivation or eradication against various pathogens should be evaluated thoroughly to understand the level of afforded disinfection. Methods commonly used in the sterilization of medical devices such as ionizing radiation, vaporized hydrogen peroxide, and ethylene oxide can provide a high level of disinfection, defined as a 6 log_10_ reduction, against bacterial spores, considered the most resistant microorganisms. CDC guidance on the decontamination and reuse of N95s [1] also includes the use of moist heat (60°C, 80% relative humidity, 15-30 min) as a possible recommendation based on literature showing preservation of fit efficiency and inactivation of H1N1 on spiked masks [2].

## Methods

### Viruses

*Bovine viral diarrhea virus* (*BVDV*), *Porcine Parvovirus* (*PPV*) *and Influenza A Virus* (*InfA*)

### Preparation of virus stocks

Virus-infected cells, cultivated in cell culture medium containing 0– 10 % (v/v) fetal bovine serum, were either frozen or thawed once prior to virus harvest. The cell debris was sedimented by centrifugation and filtration of the virus-containing cell culture supernatant through a sterile filter. The virus stock solution was stored at ≤ −60 °C in aliquots until use. The titer of the virus stock was determined according to the Spearman-Kärber method [3, 4].

### Viral titration

To determine the virus titers of the stock solution, serial three-fold dilutions were prepared with cell culture medium at a non-cytotoxic dilution. Then, 100 μL aliquots of each dilution were added to 8 wells of a 96-well-microtiter plate with indicator cells (in 100 μL cell culture medium per well). The cells were cultivated at 36.5 ± 1.0°C under 5.0 ± 1% CO_2_ in a humidified atmosphere. After a specified cultivation period, the microtiter plates were inspected microscopically for virus-induced changes in cell morphology. The viral titers were determined according to the Spearman-Kärber method.

The detection limit of a sample analyzed for the viral load is defined by the total volume incubated with the indicator cells. To improve the detection limit, a large volume of the sample was analyzed (large volume plating). Briefly, 200 µL of the diluted sample was added to a defined number of wells containing the indicator cells in 100 µL cell culture medium. The cells were cultivated for a specified incubation period. Then they were inspected microscopically for virus-induced changes in cell morphology. The indicator cells used for BVDV, PPV and InfA were BT (Taurus turbinate cell line), StNeb (Swine testis), and CRFK (Felis catus, feline epithelial cells), respectively. The corresponding growth media were Complete MEM for CRFK and StNeb cells, and Complete DMEM for BT cells.

### Viral sample preparation and spiking

The face piece or straps of several respirators were cut into equal-size pieces (n=5). Spiking at three different locations of the N95 masks were tested, namely the outer surface, the inner surface, and the straps. All spiking experiments were performed with a known concentration of virus at 5% spike ratio. Each piece was spiked with one type of virus. Three different viruses were used to challenge these masks. This process was performed at room temperature. The spiked material was placed in a sterile glass container, then left to dry in a Biosafety Cabinet for at least 1 hour.

### Viral inactivation method

The spiked samples were treated with moist heat using a circulating water bath for 30 min at 60±2°C and %80±5 humidity. The control samples (hold) were left in a biosafety cabinet until the end of the inactivation treatment. After this period, both control or treated respirator pieces were each placed in 20mL of cell/virus specific media and on an orbital shaker for at least 10 minutes to evenly disperse media across the respirator. An aliquot was removed, diluted to a non-cytotoxic (non-interfering) dilution using cell culture media and titrated accordingly. A media control was used for comparison of virus titer. Media control was performed by spiking the same amount of virus into cell/virus specific media and left for the same duration as the sample.

### Recovery and viral load assessment

The virus was considered to be recovered from the sample if a virus titer obtained from the sample was within ±1 log_10_ of the positive control. If the difference in virus titer was greater than 1 log_10_ from the titer of the positive control, the virus recovery was considered incomplete. There was no significant interference caused by the test sample if the reduction of the virus titer determined in the non-cytotoxic dilution of the test sample compared to the virus titer determined in cell culture medium was <0.5 log_10_ or within the range of the 95% confidence limit. If the difference in virus titer was > 1.0 log_10_, there was clear interference of the test sample. If the difference in virus titer was > 0.5 and ≤ 1.0 log_10_, the path forward was evaluated based on the dilution factor, the quality of the cells, and the overall CPE versus controls.

### Bacteria

*Staphylococcus aureus, Pseudomonas Aeruginosa and Acinetobacter Baumanii*

### Bacterial stock preparation

Colonies of a laboratory strain of methicillin-sensitive S. aureus (ATCC 12600) were stored at 4°C on LB agar supplemented with kanamycin (200 ug/ml) to avoid cross-contamination. Prior to antibacterial experiments, several bacterial colonies were transferred to 1.7 ml tubes (VWR) containing 1 ml of fresh LB broth and incubated at 35 ± 2 °C under 200 rpm orbital shaking until reaching turbidity of a McFarland 0.5 standard. Turbidity of the bacterial suspension was assessed using a plate-reader (BioTek Synergy H1). The stock solution had an approximate concentration of 2×10^8^ CFU/mL. *P. aeruginosa* (Rif^R^ human clinical isolate UCBPP-PA14) and *A. baumannii* (ATCC 17978) were grown on LB Lenox medium (Fisher Scientific) agar plates at 37°C. A single colony of *P. aeruginosa* and *A. baumannii* were grown in LB Lenox medium and incubated in 37°C under 200 rpm orbital shaking overnight (∼ 16 hours) in glass tubes (VWR). They were then diluted to 1:100 in 5 mL of LB before being returned to incubation in 37°C with shaking until they reached stationary phase (OD_600nm_ of slightly above 3.0). The bacterial cultures were centrifuged at 8,000 rpm for 5 minutes, and the LB supernatant was discarded. The bacterial pellet was then resuspended with PBS, using the same volume as the starting volume, yielding approximately 5×10^9^ CFU/mL.

### Bacterial sample preparation and spiking

The efficacy of moist heat decontamination was assessed using an in-vitro method adapted from an earlier paper [5]. N95 respirators (3M 1860S, St. Paul, MN) were cut into 2 cm equilateral triangles (n=4) and sterilized under UV in a biosafety cabinet for 20 minutes on each side. 100 μL of each bacterial stock suspension was transferred onto the N95 respirator samples (outer surface for *S. aureus* and inner surface for *P. aureginosa* and A. Baumanii) and air dried.

### Bacterial decontamination process

The spiked respirator pieces were placed in sterile 5 mL tubes and decontaminated for 30 minutes at 60°C and 80% relative humidity. This environment was achieved by sealing a container with saturated potassium chloride solution over which the samples were suspended in a convection oven. Positive controls were also obtained from samples that were inoculated and kept at room temperature for the duration of the decontamination procedure (hold samples).

### Recovery and bacterial load assessment

After decontamination, 5 mL of sterile phosphate buffered saline (PBS) was added, and the tubes were vortexed for 60 seconds to dislodge and resuspend bacterial inoculum. Bacterial concentration in the obtained suspension was determined using the spread-plate method outlined in reference [6]. The samples were serially diluted in PBS, and 100 μL of each dilution was spread-plated on agar plates using LB agar for *S. aureus, P. aeruginosa* isolation agar for *P. aeruginosa*, or LB Lenox medium for *A*.

### baumannii

The plates were incubated in 37°C overnight before CFUs were counted and used to calculate total CFUs from each of the conditions.

## Results

### Viruses

The control viral loads before spiking the respirators were different for the three viruses (Table 1). For PPV and BVDV, the virus concentration was maintained (<0.5 log_10_ from stock titer) when spiked and recovered from all respirator surfaces. In contrast, the infective virus concentration was decreased for InfA (<0.5 log_10_).

**Table 1.** Stock titers and recovered amount of the different viruses from respirator surfaces.

Viruses
|  |  |  |  |
| --- | --- | --- | --- |
|  | **PPV** | **BVDV** | **InfA** |
| **Control** | 8.90 | 6.83 | 7.73 |
| Recovered amount |  |  |  |
| **Outer surface** | 8.90 | 7.01 | 6.95 |
| **Inner surface** | 9.20 | 6.89 | 7.07 |
| **Straps** | 8.84 | 6.65 | 5.76 |

The control samples shown in Table 2 also include the effect of holding time for the duration of the treatment on the virus concentration on the outer surfaces of the respirators compared to the recovered samples in Table 1. The results after moist heat treatment showed that there was no effect of the treatment on the infectivity of PPV. In contrast, there was a small decrease in the infectivity of BVDV and infA was completely inactivated on the outer surfaces of the spiked respirators.

**Table 2.** Concentration of infectious virus for untreated and moist heat-treated samples. Viral inoculation was performed on the outer surface of the masks.

|  |  |  |  |
| --- | --- | --- | --- |
| **PPV (*Parvoviridae*)** |  |  |  |
| **Sample** | **Viral load, PFU/sample** | **Viral load ($\log_{10}$)** | **Reduction ($\log_{10}$)** |
| Control – viral inoculation => 30 minutes hold at RT | $2.00 \times 10^8$ | 8.30 | |
| Test – viral inoculation => decontamination for 30 minutes | $5.89 \times 10^8$ | 8.77 | **None detected** |
| **BVDV (*Flaviviridae*)** |  |  |  |
| Control – viral inoculation => 30 mi.nutes hold at RT | $4.26 \times 10^6$ | 6.63 | |
| Test – viral inoculation => decontamination for 30 minutes | $1.02 \times 10^5$ | 5.01 | **1.62** |
| **InfA (*Orthomyxoviridae*)** |  |  |  |
| Control – viral inoculation => 30 minutes hold at RT | $6.17 \times 10^5$ | 5.79 | |
| Test – viral inoculation => decontamination for 30 minutes | $\leq 27.5$ | $\leq 1.44$ | **$\geq 4.35$** |

After treatment, there was residual infective virus detected for PPV and BVDV from all tested surfaces of the respirators while there was no residual virus detected for InfA (Table 3). For PPV, the infective virus concentration was maintained on all surfaces. However, for BVDV, the log_10_ reduction of infective virus on the straps was higher than that for the inner and outer surfaces.

**Table 3.**
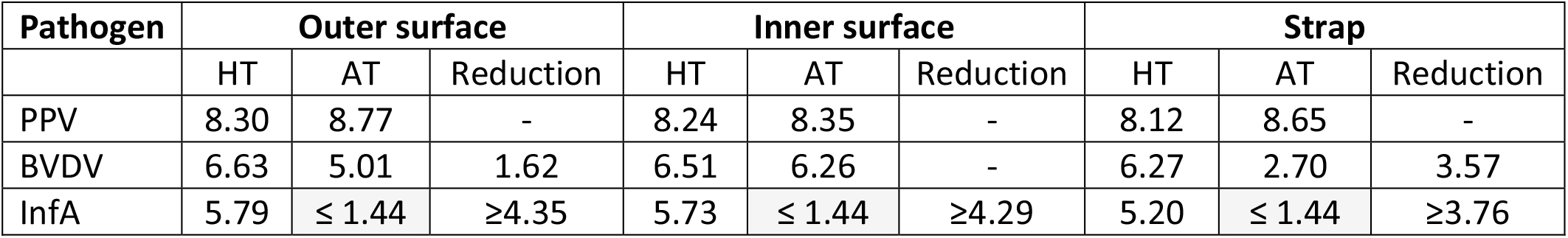
Comparison of different surfaces of absorption for inactivation of pathogens. Concentrations are reported in log_10_. HT: Hold titer, AT: After treatment. The highlighted cells (gray) indicate no residual virus was detected.

|  |  |  |  |  |  |  |  |  |  |
| --- | --- | --- | --- | --- | --- | --- | --- | --- | --- |
| Pathogen | Outer surface |  |  | Inner surface |  |  | Strap |  |  |
|  | HT | AT | Reduction | HT | AT | Reduction | HT | AT | Reduction |
| PPV | 8.30 | 8.77 | - | 8.24 | 8.35 | - | 8.12 | 8.65 | - |
| BVDV | 6.63 | 5.01 | 1.62 | 6.51 | 6.26 | - | 6.27 | 2.70 | 3.57 |
| InfA | 5.79 | $\leq 1.44$ | $\geq 4.35$ | 5.73 | $\leq 1.44$ | $\geq 4.29$ | 5.20 | $\leq 1.44$ | $\geq 3.76$ |

**Table 4.**
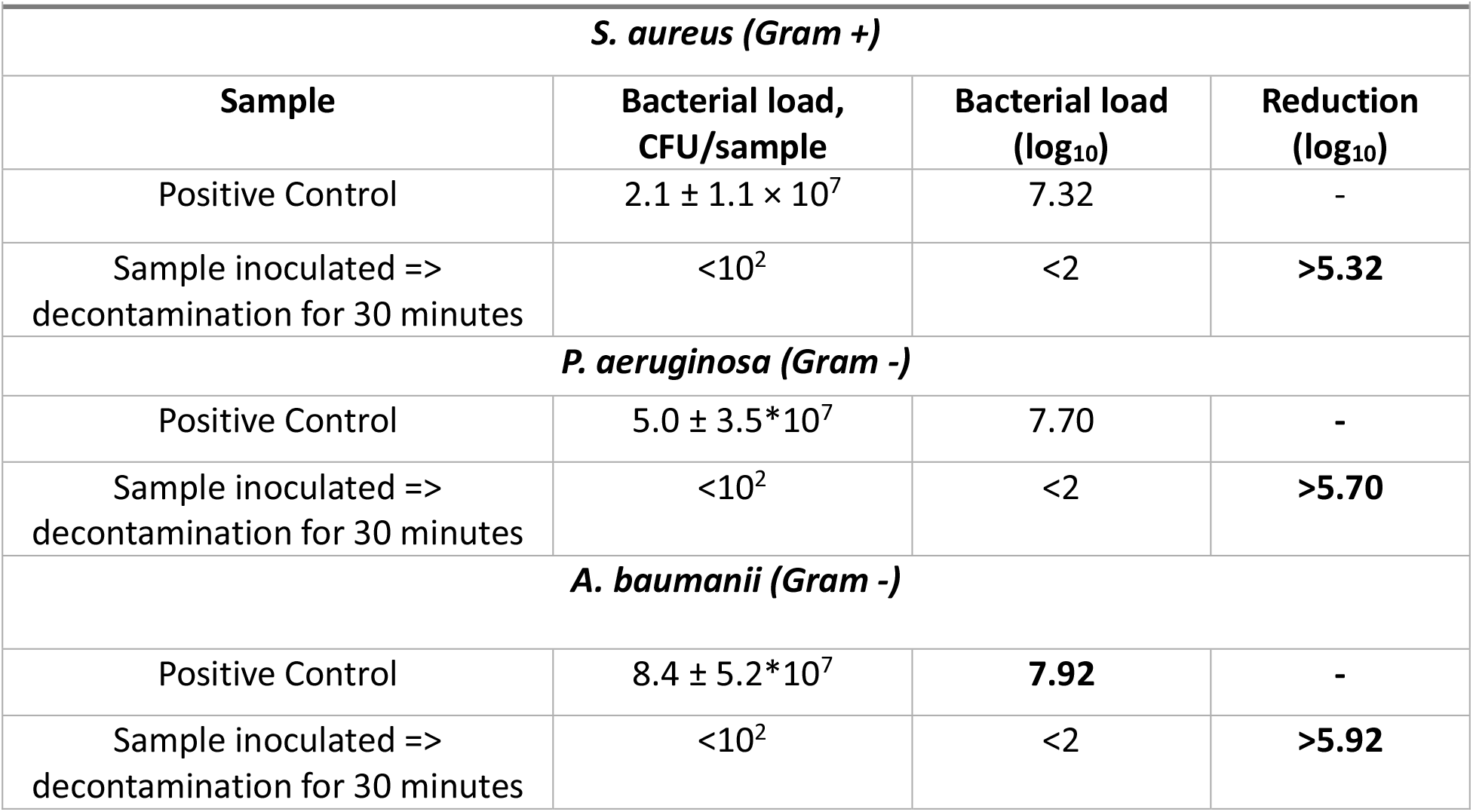
Concentration of bacteria for untreated and moist heat-treated samples.

The detection limit of bacterial assays was 10^2^ CFU (1 CFU/plate). No colonies grew on any of the plates for the treated samples or the negative controls. Based on the concentration of the control, the reduction of the bacteria concentration ranged from 5.32 to 5.92.

## Discussion

Some standard techniques for the sterilization of medical devices and tools such as vapor hydrogen peroxide have been shown to be effective against SARS-CoV2, as well as more resistant pathogens such as *Geobacillus stearothermophilus* spores [7]. However, in the absence of spore testing or for the methods that are not expected to be effective against the entire spectrum of pathogen resistance (Figure 1), it is desirable to evaluate the level of disinfection provided.

**Figure 1.**
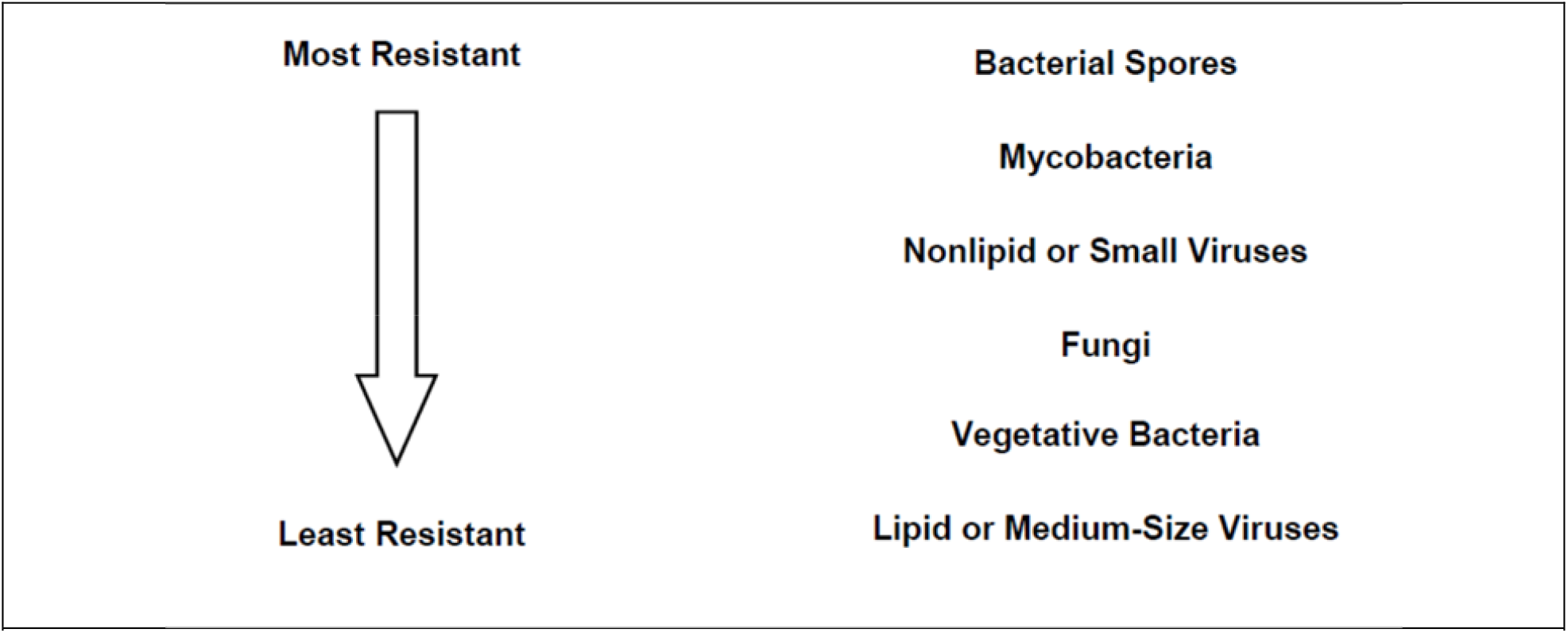
Schematic adapted from FDA guidance entitled ‘Enforcement policy on sterilizers, disinfectant devices, and air purifiers during the coronavirus disease 2019 (COVID-19) public health emergency’ (March 2020).

The obtained results showed the limits of efficacy of moist heat decontamination against various pathogens. Moist heat decontamination under the conditions studied here yielded at least a 5.3 log reduction with no residual colonies against the vegetative bacteria *S. aureus, P. aeruginosa* and *A. Baumannii*. On the other hand, the method’s efficacy against the tested viruses varied greatly; it was effective against InfA, modestly effective against BVDV, and not effective at all against PPV.

Vegetative bacteria are slightly more resistant to disinfection than medium-size lipid or enveloped viruses such as SARS-CoV2. *S. aureus* is a common microorganism found on human skin, and it is one of the most common causative pathogens of both community-and hospital-acquired infections [8]. *S. aureus* is often associated with a variety of infectious conditions such as bacteremia, skin and soft tissue infections, osteomyelitis, septic arthritis, urinary tract infections among others [9]. Treatment of *S. aureus* associated infections can be challenging due to the emergence of antibiotic resistant strains, such as methicillin-resistant S. aureus (MRSA) [10, 11]. *P. aeruginosa* and *A. baumannii* are recalcitrant ESKAPE Gram-negative bacteria that exemplify current highly problematic clinical pathogens causing serious infections in hospital and community settings, due to the rapidly acquired multidrug resistance (MDR), the formation of biofilms and antibiotic tolerant/persister (AT/P) cells [12]. They are globally prevalent and, more importantly, the mortality associated with MDR resistant infections caused by these bacteria is especially high (39 and 80%, respectively; 13 and 14]. *A. baumanni* survives for prolonged periods under a wide range of environmental conditions and causes outbreaks of infection and health care-associated infections, including bacteremia, pneumonia, meningitis, urinary tract infection, and wound infection. InfA and BVDV are enveloped, single strand RNA viruses, similar to SARS-CoV2. However, BVDV is likely harder to eradicate due to its small size (40-60 nm) and has been shown to persist for up to 3 weeks in farm slurry at 5° C [15]. PPV is a small, non-enveloped, DNA virus (18-24 nm), which is known to be more resistant to disinfection than enveloped viruses and bacteria (Figure 1).

The FDA guidance document “Enforcement Policy for Face Masks and Respirators During the Coronavirus Disease (COVID-19) Public Health Emergency (Revised)” (April 2020) recommends that a candidate decontamination method for respirators demonstrate high-level disinfection using a ≥ 6-log_10_ reduction of bacterial spores and a > 3 log_10_ reduction in viral infectivity where viral titers will allow. In a different guidance document entitled ‘Enforcement Policy for Sterilizers, Disinfectant Devices, and Air Purifiers During the Coronavirus Disease 2019 (COVID-19) Public Health Emergency (March 2020), three levels of disinfection are clearly defined based on the activity of the disinfection method against different pathogens. According to these definitions, a ‘low’ level of disinfection could be obtained by the demonstration of 6- log_10_ reduction of each of the typical vegetative organisms *Pseudomonas aeruginosa, Staphylococcus aureus, Escherichia coli*, and representatives of the *Klebsiella* and *Enterobacter genus*. This level of disinfection is described as ‘a lethal process utilizing an agent that kills vegetative forms of bacteria, some fungi, and lipid viruses’. According to this guidance and definitions, moist heat decontamination under the tested conditions here is likely to provide low level disinfection.

The factors that are considered for the effective reuse of these respirators aside from viral inactivation are the fit, and filter efficiency. Moist heat decontamination under these conditions has been shown previously to preserve fit and filter efficiency for N95 respirators [2]. One of the factors to be considered while evaluating the merits of different decontamination methods is the number of possible decontamination cycles before the fit, filter efficiency or efficacy of bioburden reduction is compromised. Other factors are the use patterns of the respirators in between decontamination treatments and whether the respirators are returned to the original users after decontamination. These factors may affect the performance and the risk of failure of these decontaminated respirators. There is little information on the effect of multiple decontamination cycles using moist heat decontamination under these conditions, but a recent non-peer reviewed study suggests that the filter efficiency may be maintained up to 20 cycles [16].

While we present results for viral and bacterial bioburden reduction of this decontamination method for one type of N95 respirator, there are many other considerations such as other technical, financial, and logistical ones for any medical institution to decide on a decontamination method, if any, to utilize for this emergency use situation.

## Data Availability

The data obtained is largely presented in the manuscript. Any other details which may not be directly presented may be requested form the authors.

